# DNA methylation and general psychopathology in childhood: An epigenome-wide meta-analysis from the PACE consortium

**DOI:** 10.1101/2022.01.26.22269579

**Authors:** Jolien Rijlaarsdam, Marta Cosin, Laura Schellhas, Sarina Abrishamcar, Anni Malmberg, Alexander Neumann, Janine F. Felix, Jordi Sunyer, Kristine B. Gutzkow, Regina Grazuleviciene, John Wright, Mariza Kampouri, Heather J. Zar, Dan J. Stein, Kati Heinonen, Katri Räikkönen, Jari Lahti, Anke Huels, Doretta Caramaschi, Silvia Alemany, Charlotte A. M. Cecil

**Author notes:** Contributed equally (Jolien Rijlaarsdam, Marta Cosin, Laura Schellhas, Doretta Caramaschi, Silvia Alemany, Charlotte A. M. Cecil).

## Abstract

The general psychopathology factor (GPF) has been proposed as a way to capture variance shared between psychiatric symptoms. Despite a growing body of evidence showing both genetic and environmental influences on GPF, the biological mechanisms underlying these influences remain unclear. In the current study, we conducted epigenome-wide meta-analyses to identify both probe- and region-level associations of DNA methylation (DNAm) with school-age general psychopathology in six cohorts from the Pregnancy And Childhood Epigenetics (PACE) Consortium. DNAm was examined both at birth (cord blood; prospective analysis) and during school-age (peripheral whole blood; cross-sectional analysis) in total samples of N=2,178 and N=2,190, respectively. At school-age, we identified one probe (cg11945228) located in the Bromodomain-containing protein 2 gene (*BRD2*) that negatively associated with GPF (*p*=8.58×10^−8^). We also identified a significant DMR at school-age (*p*=1.63×10^−8^), implicating the SHC Adaptor Protein 4 (*SHC4*) gene that has been previously implicated in multiple types of psychiatric disorders in adulthood, including obsessive compulsive disorder and major depressive disorder. In contrast, no prospective associations were identified with DNAm at birth. Taken together, results of this study revealed some evidence of an association between DNAm at school-age and GPF. Future research with larger samples is needed to further assess DNAm variation associated with GPF.

## Introduction

Psychiatric disorders or symptoms co-occur more often than would be expected by chance alone.^1, 2^ In light of the negative clinical and functional outcomes associated with psychiatric co-occurrence,^3, 4^ it is important to identify early indicators of risk and underlying biological mechanisms. There is accumulating evidence that, as early as in childhood, the shared variance between psychiatric disorders or symptoms may be usefully represented by a general psychopathology factor (GPF).^5-8^ This GPF in childhood has been found to show temporal stability^6^ and to predict long-term functional and psychiatric outcomes in adolescence throughout adulthood.^9, 10^ Although previous research has found evidence for both genetic and environmental influences on GPF,^7, 11-16^ the biological mechanisms underlying these influences remain unclear.

One of the ways by which genetic and environmental factors might contribute to disease susceptibility is through epigenetic mechanisms that regulate gene expression, such as DNA methylation (DNAm).^17^ Studies have shown that variation in DNAm is influenced by a dynamic interplay of genetic and environmental factors.^18^ In turn, alterations in DNAm patterns across the genome have been found to associate with a wide range of child and adult mental health outcomes, such as conduct problems, attention deficit hyperactivity disorder (ADHD) symptoms, major depressive disorder (MDD), and schizophrenia.^19-22^ Despite a growing body of research implicating an involvement of DNAm in individual mental health outcomes, much less work has focused on the relationship between DNAm and general psychopathology.^23^ To the best of our knowledge, only one study examined the association between genome-wide DNAm patterns and GPF in childhood. In this study, data were analyzed cross-sectionally in one cohort, focusing on wider biological networks (so called ‘modules’) of co-methylated DNAm probes across the genome.^23^ As such, we still lack knowledge on how GPF relates to single DNAm probes and the extent to which associations vary across time (i.e., both cross-sectional and prospective associations). Large multi-cohort epigenome-wide studies, which allow for increased power and generalizability, are needed to improve our understanding of the biological mechanisms underlying shared variance across mental health problems.

We conducted epigenome-wide meta-analyses to investigate both probe-level and region-based associations of DNAm with school-age GPF in the Pregnancy And Childhood Epigenetics (PACE) Consortium. Because it is unclear at which time point differential DNAm may be most relevant to GPF, we examined DNAm both at birth (cord blood; prospective study; pre-symptom manifestation) and at school-age (peripheral whole blood; cross-sectional study) in pooled samples of N=2,178 and N=2,190 children, respectively.

## Methods

### Participants

The prospective analyses included four cohorts from PACE, using complete data on DNAm at birth, general psychopathology in childhood and covariates: the Avon Longitudinal Study of Parents and Children (ALSPAC), Drakenstein Child Health Study (DCHS), Generation R (GENR), and INfancia y Medio Ambiente (INMA). These cohorts have a combined sample size of 2,178 (see **Table 1**). All prospective cohorts included participants of European ancestry, except for DCHS, which included participants of predominantly Black African ancestry or mixed ancestry. See **Supplementary Methods** for full cohort descriptions.

**Table 1.**
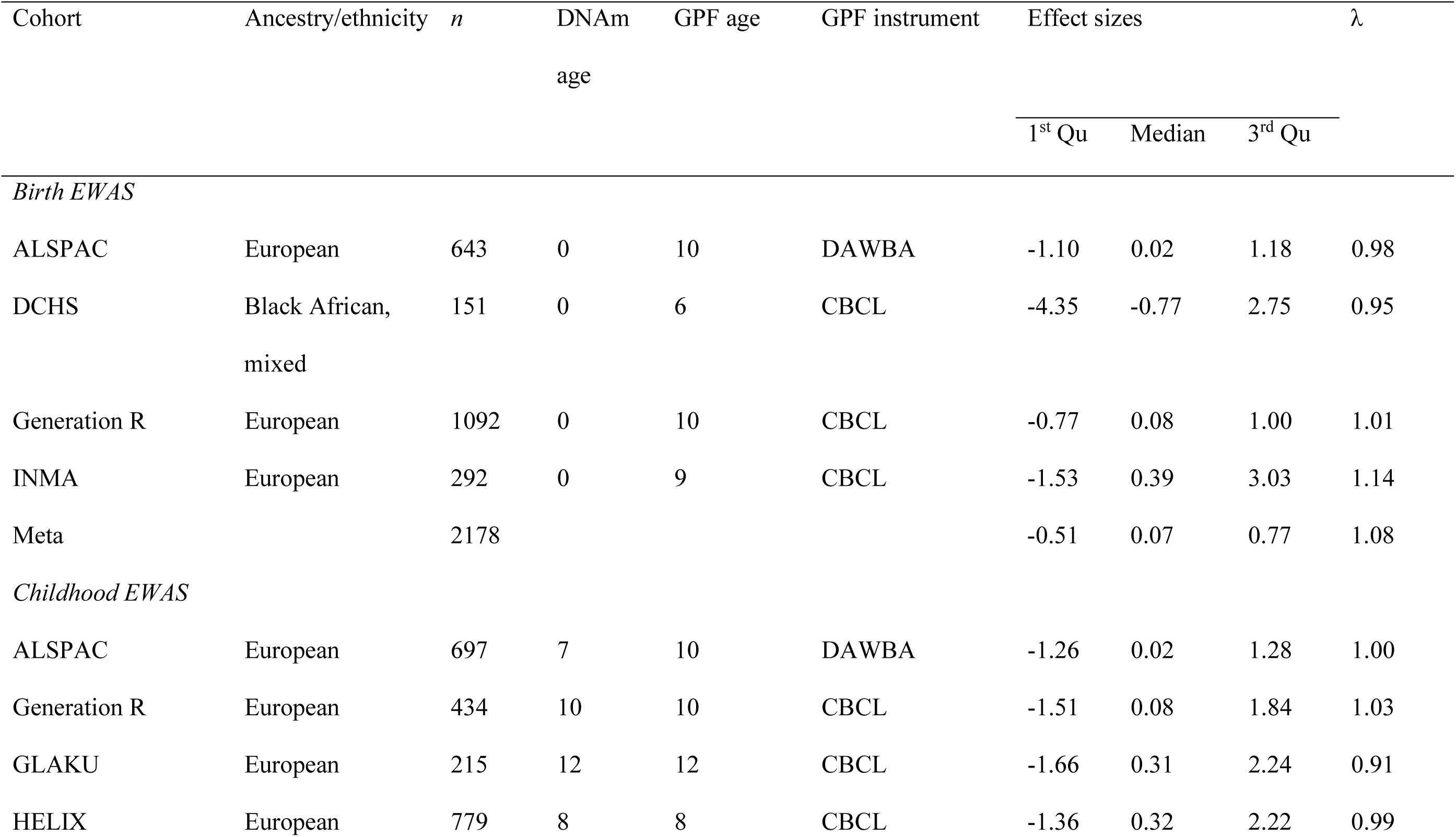
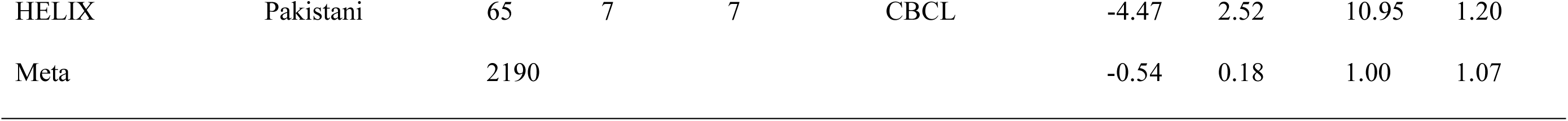
Cohort characteristics

The cross-sectional analyses included four cohorts from the PACE consortium, using complete data on DNAm and general psychopathology in childhood, as well as covariates; ALSPAC, GENR, Glycyrrhizin in Licorice (GLAKU), and Human Early Life Exposome (HELIX; including six jointly analyzed sub cohorts). These cohorts have a combined sample size of 2,190 (see **Table 1**). All cross-sectional cohorts included participants of European ancestry, except for HELIX, which included participants of European ancestry and participants with a Pakistani background living in the United Kingdom, which were analyzed as separate cohorts in our meta-analysis.

### Measures

#### DNA methylation

DNAm was assessed with the Illumina Infinium HumanMethylation450 (ALSPAC, DCHS, GENR, HELIX, INMA) or the Infinium HumanMethylationEPIC (DCHS, GLAKU) BeadChip assays in cord blood and in peripheral whole blood at ages 7-12 years. The cohorts performed sample processing, quality control (QC) and normalization based on their preferred protocols as described in the **Supplementary methods**. We used normalized, untransformed beta values, ranging from 0 (fully unmethylated) to 1 (fully methylated). Methylation levels that fell outside of the lower quartile minus 3 × interquartile or upper quartile plus 3 × interquartile range were removed.

We excluded probes with a call rate <90%, control probes, and probes that mapped to X/Y chromosomes. Following Zhou et al.,^24^ we further excluded probes with poor base pairing quality (lower than 40 on 0-60 scale), probes with non-unique 30bp 3’-subsequence (with cross-hybridizing problems), Infinium II probes with SNPs of global MAF over 1% affecting the extension base, and probes with a SNP in the extension base that causes a color channel switch from the official annotation. We also excluded a subset of probes (*n*=69) that have shown to be unreliable in a recent comparison of the Illumina 450K and EPIC BeadChips.^25^ At the meta-analysis level, we excluded probes which were available in <50% of the cohorts and <50% of the participants. After QC, 404,017 probes remained at birth and 413,497 probes remained at school-age.

#### General psychopathology factor

Mental health symptoms were assessed when children were aged 6-10 years, depending on the cohort. Parent-rated instruments were used, including (i) the Child Behavior Checklist 6-18 (CBCL/6-18) in DCHS, GENR, GLAKU, HELIX, and INMA, and (ii) the Development and Well-being Assessment (DAWBA) in ALSPAC. Instruments are described in the Supplementary methods.

#### Covariates

We adjusted for the following potential confounders: child sex, gestational age at birth, child age at the assessment of outcome, maternal age, maternal educational level, prenatal maternal smoking status, cell-type proportions estimated using standard algorithms for DNAm at birth^26^ or childhood,^27^ ancestry (depending on the specific cohort), and technical covariates (e.g., batch) (see **Supplementary methods**).

### Statistical analyses

#### General psychopathology factor

We used confirmatory factor analysis (CFA) to fit a general psychopathology model in the full samples with mental health data available (see **Supplementary Information 1**). Each cohort ran the CFA according to a predefined script, using the Lavaan statistical package^28^ in R (https://www.r-project.org/). GPF scores were extracted, winsorized at +/- 3SD, and standardized.

#### Cohort-specific EWAS

Each cohort ran the EWAS according to a predefined analysis plan, using robust linear regression (rlm; *MASS* R-package) to account for potential heteroscedasticity and non-normality. Cohorts excluded all multiple births and chose one random sibling per non-twin sibling pair.

#### Meta-analysis

The cohort-specific results were meta-analyzed at Erasmus MC Rotterdam. A shadow meta-analysis was conducted independently at the Barcelona Institute for Global Health. We performed an inverse-variance weighted fixed effects approach using R and METAL.^29^ Probes were annotated using *meffil*.^30^ Genome-wide significance was defined at the Bonferroni-corrected threshold of *p*< 1×10^−7^, and suggestive significance at *p*< 1×10^−5^. We included *p*< 1×10^−4^ specifically for pathway and enrichment analyses to allow a sufficient number of genes to be included.

We ran two sensitivity meta-analyses. First, we included only cohorts of predominantly European ancestry to check if the results of the main analysis were influenced by ancestry. Second, we performed leave-one-out meta-analyses for hits showing genome-wide significant associations with GPF to ensure that associations were not driven by a single cohort.

#### Differentially methylated regions

Differentially methylated regions (DMRs) were identified using the dmrff package^31^ in R. This method first identifies candidate DMRs by screening the meta-level EWAS results for genomic regions each covered by a sequence of CpG sites with EWAS effects in the same direction, EWAS *p*-values <0.05, and <500bp gaps between consecutive CpG sites. Then, summary statistics are calculated for each candidate DMR within each of the cohorts by meta-analyzing the cohort-level EWAS summary statistics of the CpG sites in the region. Meta-analysis is performed by a variation of inverse weighted fixed effects meta-analysis that accounts for non-independence between CpG sites. Finally, for each candidate DMR, the summary statistics from each cohort are meta-analyzed to obtain a cross-cohort meta-analyzed DMR statistic and *p*-value.

#### Follow-up analyses

To characterize potential genetic and environmental influences of individual probes showing genome-wide or suggestive significance, we first used a heritability tool quantifying additive genetic influences as opposed to shared and non-shared environmental influences on DNAm, based on data from monozygotic and dizygotic twins.^32^ We then used the GoDMC database (http://mqtldb.godmc.org.uk/) as a more specific tool for identifying genetic influences on DNAm levels via mQTL mapping. Third, we characterized cross-tissue correspondence of DNAm using the blood-brain concordance tool^33^ based on postmortem data from 122 individuals with DNAm from whole blood and four brain regions. Finally, to assess whether methylation levels of CpGs were associated with the expression levels of nearby genes in child blood, we consulted the HELIX Expression Quantitative Trait Methylation (eQTM) catalogue (https://helixomics.isglobal.org/).

To identify broader pathways and enrichment for molecular functions, we used the gene ontology (GO-biological processes, GO-molecular functions and GO-cellular components), the Kyoto Encyclopedia of Genes and Genomes (KEGG) pathway, and the Molecular Signature Database (MSigDB) enrichment methods from the missMethyl R package,^34^ as implemented in the Functional Enrichment module of the EASIER R package.^35^ We ran GWAS enrichment analyses for EWAS using the GenomicRanges Package,^36^ to identify genomic regions of EWAS hits (*p*< 1×10^−4^) that overlapped with the 361 genome-wide significant loci previously reported in GWASs on general psychopathology,^16^ schizophrenia,^37^ or neuroticism^38^ (0.5Mb window centered to the genomic locus indicated in the original studies).

## Results

### General psychopathology factor

All mental health subscales had significant loadings on the general factor across all cohorts, with all loadings >0.30. For full details on the GPF loadings, correlations, and model fit, **see Supplementary Table 1**. In line with previous research,^5, 7, 14^ GPF consistently negatively correlated with child cognition (see **Supplementary methods**) across the cohorts (mean *r*=-0.12, range=-0.08 to -0.13).

### Epigenome-wide meta-analysis

Descriptive statistics across the cohorts are shown in **Supplementary Table 2**. We prospectively examined associations of DNAm at birth (*n*=2,178) at 404,017 CpG sites with GPF at school-age. There was no evidence of genomic inflation in the cohort-specific EWASs (range λ=0.95-1.14), nor in the meta-analysis (λ=1.08, see also **Figure 1**). As can be seen in **Figure 2**, no CpG reached genome-wide significance at *p*< 1×10^−7^, with four CpGs showing *p*< 1×10^−5^ (see **Table 2**). For the top hit (cg02084087), annotated to *TNFRSF25* (TNF Receptor Superfamily Member 25), a 10-point increase in percentage methylation was related to a 0.43 SD increase in general psychopathology symptoms (*p*=5.54×10^−6^).

**Table 2.** DNA methylation at birth and general psychopathology: meta-analytic associations with *p* < 1×10^−5^

| CpG | Gene | Chr | Position | *n* | B | SE | *p* | Direction | *I*<sup>2</sup> | Heterogeneity<br>*p*-value |
| --- | --- | --- | --- | --- | --- | --- | --- | --- | --- | --- |
| cg02084087 | *TNFRSF25* | chr1 | 6526049 | 2175 | 4.31 | 0.95 | $5.54 \times 10^{-6}$ | ++++ | 46.2 | 0.13 |
| cg11777523 | *GPR148* | chr2 | 131485418 | 2019 | 9.26 | 2.07 | $7.80 \times 10^{-6}$ | +?++ | 23.6 | 0.27 |
| cg14358879 | *SLC8A3* | chr14 | 70655920 | 2174 | -7.82 | 1.75 | $8.20 \times 10^{-6}$ | ---- | 33.7 | 0.21 |
| cg09437808 | - | chr5 | 176107069 | 2177 | 2.46 | 0.55 | $8.96 \times 10^{-6}$ | ++++ | 0 | 0.47 |

Chr = chromosome, *n* = number of participants, SE = standard error, Direction = direction of the effect per study (ALSPAC, DCHS, GenR, INMA) in alphabetical order (+ = positive direction, - = negative direction, ? = not present); *I*<sup>2</sup> = heterogeneity statistic reflecting the variation attributable to heterogeneity across studies (high values suggest high heterogeneity)
*Note.* Effect estimates (B) represent the SD increase in GPF for each increase of 100% in DNAm.

**Table 3.** DNA methylation at school-age and general psychopathology: meta-analytic associations with *p* < 1 ×10^−5^

| CpG | Gene | Chr | Position | *n* | B | SE | *p* | Direction | I<sup>2</sup> | Heterogeneity<br>*p*-value |
| --- | --- | --- | --- | --- | --- | --- | --- | --- | --- | --- |
| cg11945228 | *BRD2* | chr6 | 32940368 | 2173 | -37.00 | 6.91 | 8.58x10<sup>-8</sup> | ----+ | 0 | 0.53 |
| cg18862005 | - | chr2 | 177940863 | 1972 | 3.78 | 0.72 | 1.78x10<sup>-7</sup> | ++?++ | 0 | 0.64 |
| cg22691524 | - | chr3 | 185300576 | 2180 | 7.84 | 1.52 | 2.54x10<sup>-7</sup> | +++++ | 0 | 0.47 |
| cg00719568 | *MOV10* | chr1 | 113239645 | 2184 | 4.20 | 0.83 | 3.62x10<sup>-7</sup> | +++++ | 0 | 0.67 |
| cg09040034 | *KIFC1* | chr6 | 33362567 | 1966 | 4.89 | 1.01 | 1.25x10<sup>-6</sup> | ++?+- | 40 | 0.17 |
| cg24514921 | *VPS54* | chr2 | 64246311 | 2184 | 12.17 | 2.51 | 1.27x10<sup>-6</sup> | ++++- | 0 | 0.45 |
| cg25182716 | - | chr20 | 13622875 | 2178 | 6.42 | 1.36 | 2.52x10<sup>-6</sup> | +++++ | 0 | 0.76 |
| cg08514304 | *TAOK2* | chr16 | 29994437 | 2175 | 6.88 | 1.48 | 3.36x10<sup>-6</sup> | +++++ | 0 | 0.59 |
| cg00470277 | - | chr7 | 2669915 | 2185 | 4.55 | 0.98 | 3.62x10<sup>-6</sup> | +++++ | 0 | 0.99 |
| cg26353764 | *WDR20* | chr14 | 102660055 | 2187 | 6.82 | 1.48 | 3.86x10<sup>-6</sup> | +++++ | 43.7 | 0.13 |
| cg27009703 | *HOXA9* | chr7 | 27204894 | 2121 | -8.58 | 1.86 | 3.93x10<sup>-6</sup> | ----- | 0 | 0.52 |
| cg12492087 | *ZFP106* | chr15 | 42749885 | 2178 | 6.39 | 1.39 | 4.01x10<sup>-6</sup> | +++++ | 0 | 0.92 |
| cg18436008 | - | chr10 | 80535327 | 2176 | 5.48 | 1.19 | 4.31x10<sup>-6</sup> | +++++ | 0 | 0.56 |
| cg17281031 | - | chr12 | 128223216 | 2186 | 4.50 | 0.98 | 4.73x10<sup>-6</sup> | +++++ | 14.6 | 0.32 |
| cg08327106 | *RALYL* | chr8 | 85094842 | 2175 | 8.60 | 1.89 | 5.60x10<sup>-6</sup> | +++++ | 0 | 0.66 |
| cg11236841 | - | chr4 | 35567978 | 2175 | 4.67 | 1.03 | 6.08x10<sup>-6</sup> | +++++ | 54.3 | 0.07 |
| cg21525176 | *LHFPL2* | chr5 | 77906752 | 2182 | 6.09 | 1.36 | 7.19x10<sup>-6</sup> | ++-++ | 59.2 | 0.04 |
| cg26420013 | *NSUN2;SRD5A1* | chr5 | 6632020 | 2189 | 2.44 | 0.55 | 8.03x10<sup>-6</sup> | +++++ | 56.1 | 0.06 |
| cg17087232 | *MAN2C1* | chr15 | 75651821 | 2187 | 3.45 | 0.78 | 9.00x10<sup>-6</sup> | +++++ | 0 | 0.98 |
| cg16360861 | *RAI14* | chr5 | 34684597 | 2175 | 5.12 | 1.16 | 9.36x10<sup>-6</sup> | +++++ | 0 | 0.53 |
| cg00737264 | *SMOC2* | chr6 | 169049498 | 2188 | 1.88 | 0.42 | 9.40x10<sup>-6</sup> | + -+++ | 41.3 | 0.15 |

Chr = chromosome, *n* = number of participants, SE = standard error, Direction = direction of the effect per study (ALSPAC, GenR, GLAKU, HELIX, HELIX Pakistani) in alphabetical order (+ = positive direction, - = negative direction, ? = not present); $I^2$ = heterogeneity statistic reflecting the variation attributable to heterogeneity across studies (high values suggest high heterogeneity)
*Note.* Effect estimates (B) represent the SD increase in GPF for each increase of 100% in DNAm.

**Fig. 1.**
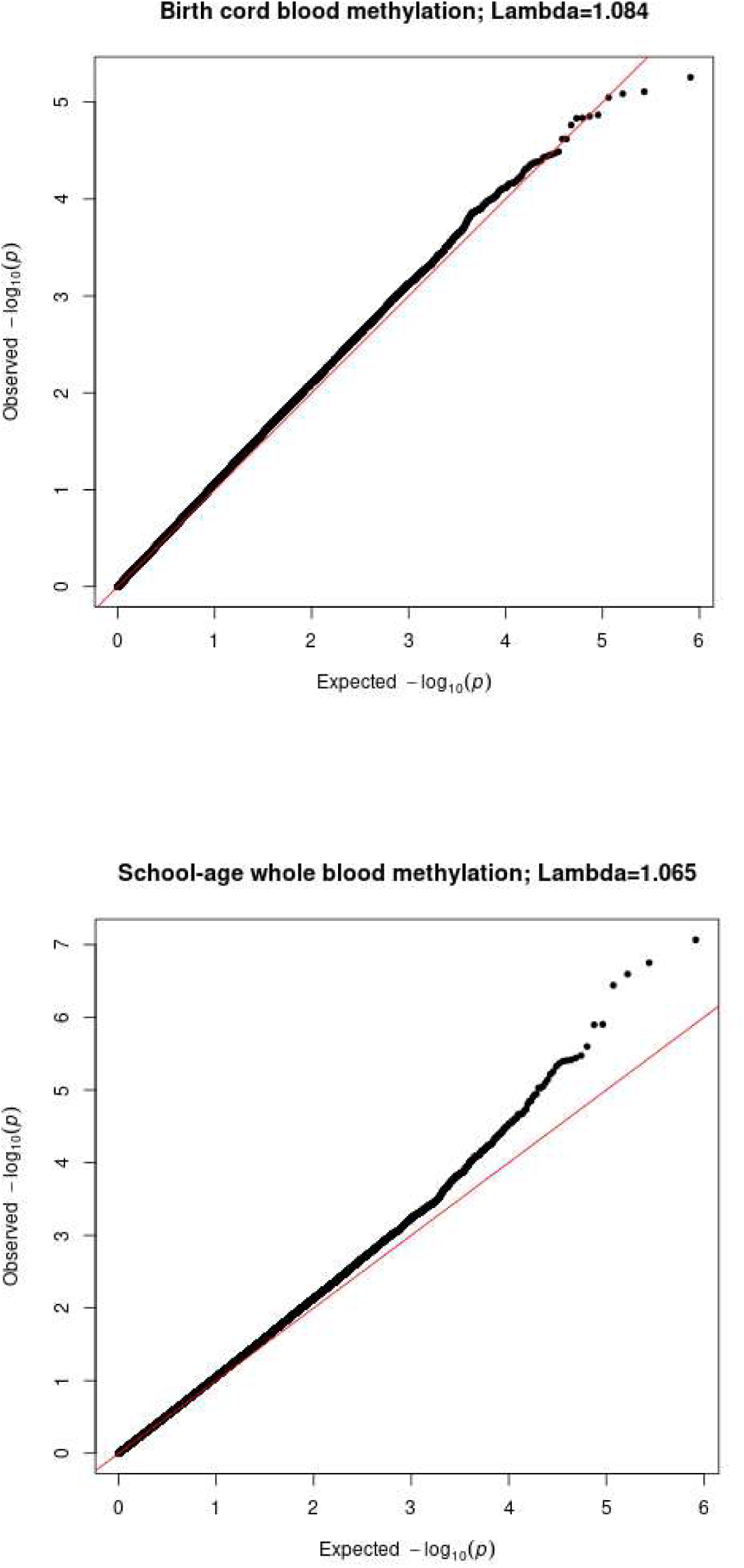
Quantile-quantile plot of the meta-analytic associations of DNA methylation at birth and DNA methylation at school-age with general psychopathology. The diagonal line represents the distribution of the expected *p*-values under the null. Points above the diagonal refer to *p*-values that are lower than expected.

**Fig. 2.**
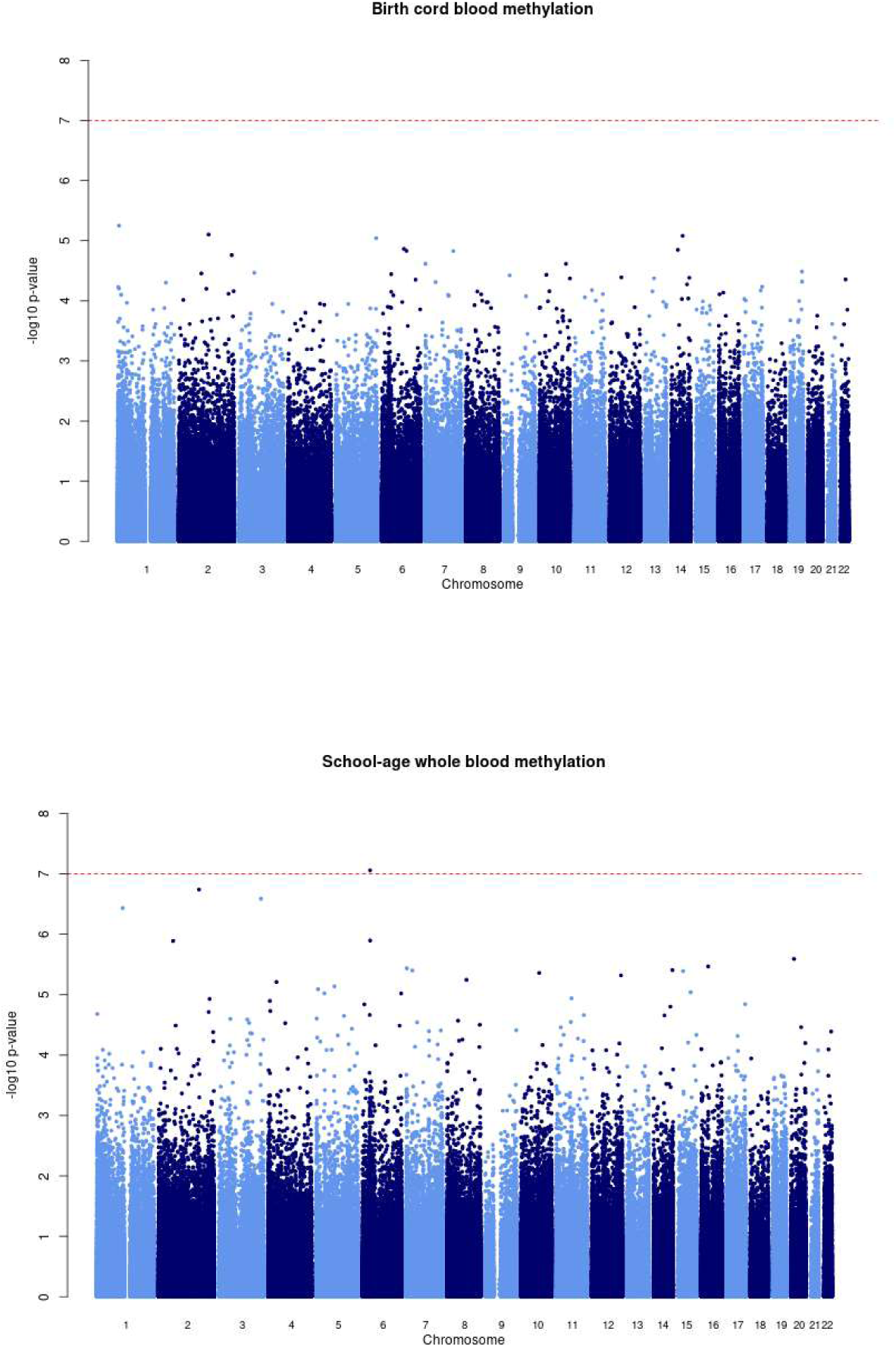
Manhattan plot of –log_10_ *p*-values versus CpG position (base pair and chromosome) showing meta-analytic associations of DNA methylation at birth and DNA methylation at school-age with general psychopathology. The red line indicates genome-wide significance (*p* < 1 ×10^−7^).

In the cross-sectional meta-analysis of DNAm at school-age (*n*=2,190) at 413,497 sites, one CpG reached genome-wide significance (see **Figure 2**). For this CpG probe (cg11945228), mapped to *BRD2* (Bromodomain-containing protein 2 gene), a 10-point increase in percentage methylation was related to a 3.70 SD decrease in general psychopathology symptoms (*p*=8.58×10^−8^). Twenty other CpGs showed *p*< 1×10^−5^. These 21 top hits identified at school-age did not overlap with the ones observed at birth. Furthermore, as shown in **Supplementary Table 3**, the significant hit identified at school-age did not reach nominal significance (*p* <0.05) at birth (B=5.28, SE=3.76, *p*=0.16). Nominally significant probes identified in childhood correlated at *r*=0.004, *p*=0.55 (*n*=23,764) with respective probes at birth.

### Sensitivity analyses

Restricting the meta-analysis to children with European ancestry did not change the overall pattern of results for both prospective (*n*=2,027) and cross-sectional (*n*=2,125) studies, as evidenced by cross-meta-analysis correlations of effect estimates (*r*_prospective_=0.99, *r*_cross-sectional_=0.99) and consistent directions (95% and 96%, respectively) of effect estimates. The top hit identified at school-age remained genome-wide significant (B=-38.02, SE=6.95, *p*=4.47×10^−8^).

Leave-one-out meta-analyses showed that the significant top hit identified during childhood (cg11945228) was robust to excluding all individual cohorts, except GenR (for a leave-one-out plot, see **Supplementary Figure 1**). Furthermore, when looking at the cohort-level EWAS results, the cross-sectional association between cg11945228 and GPF was statistically significant in GenR (B=-41.87, SE=7.78, *p*=7.39×10^−8^) but not in the other cohorts (all *p* >0.25, see **Supplementary Figure 2** for a forest plot).

### Differentially methylated regions

In the prospective analyses, there was no evidence of DMRs at birth associated with GPF. In the cross-sectional analyses, one DMR at childhood was associated with GPF (estimate=10166.54, SE=1800.19, *p*=1.63×10^−8^). As shown in **Supplementary Table 4**, this DMR included 6 CpGs mapped to the *SHC Adaptor Protein 4* gene (*SHC4*) at chromosome 15, all located in the gene body.

### Follow-up analyses

All probes showing significant or suggestive associations with DNAm had twin heritability estimates available, showing mean additive genetic influences of *r*_birth_=0.16 and *r*_childhood_=9.44×10^−2^ (**see Supplementary Table 5**).

Of the four suggestive probes identified at birth, three were associated with at least one known methylation quantitative trait locus (mQTL) (**see Supplementary Table 5**) and one showed a concordant DNAm pattern (*r* >0.28, *p* <0.01) across blood versus the prefrontal cortex, entorhinal cortex, and the superior temporal gyrus (**see Supplementary Table 6**).

The genome-wide significant probe identified during childhood (cg11945228) was unrelated to known mQTLs and showed non-significant correlations between blood versus brain DNAm. Of the 20 suggestive probes identified in childhood, ten were associated with at least one known mQTL (**see Supplementary Table 5**) and four showed a significant correlation between blood versus brain DNAm (*r* > 0.25, *p* <0.04; **see Supplementary Table 6**). None of the suggestive probes identified at birth or childhood showed links to an eQTM.

The six probes in the DMR at childhood showed mean additive genetic influences of *r*=0.007. None of these probes related to mQTLs or eQTMs. One probe (cg05867423) showed a concordant DNAm pattern (*r* =-0.36, *p* =0.002) across blood versus the prefrontal cortex.

GO, KEGG, and MSigDB analyses revealed no significantly enriched common biological processes, cellular components, molecular functions or pathways for the genes mapped to the probes at *p*< 1×10^−4^ in the meta-analyses at birth (*n*=56) and during childhood (*n*=104) (see **Supplementary Tables 7a-f**).

Results of the GWAS enrichment analyses for EWAS are presented in **Supplementary Table 8**. Of the 56 probes at *p*< 1×10^−4^ in the prospective EWAS meta-analysis, six overlapped with genomic loci previously linked to general psychopathology, schizophrenia, or neuroticism based on GWASs.^16, 37, 38^ Of the 104 probes at *p*< 1×10^−4^ in our cross-sectional EWAS meta-analysis, 13 (12.5%) overlapped with genomic loci previously linked to these psychiatric outcomes. Most notably, this cross-sectional enrichment analysis prioritized cg08514304 (*TAOK2*), which was among the top 10 suggestive hits identified in our cross-sectional EWAS meta-analysis and showed a consistent direction of effect in all cohorts.

## Discussion

We conducted the largest epigenome-wide meta-analysis of GPF in childhood, using DNAm assessments at two different time points (birth and childhood). The analyses revealed little evidence for probe-specific associations between DNAm in peripheral blood and GPF. However, we did identify a significant DMR in childhood, implicating a gene with pleiotropic effects.

On the basis of probe-level genome-wide meta-analyses, we found that lower DNA methylation at cg11945228 at school-age was significantly associated with higher levels of GPF. Cg11945228 is located within the *BRD2* gene, a BET (bromodomains and extra terminal domain) family chromatin adaptor that controls the transcription of a wide range of pro-inflammatory genes^39^ and is involved in neural tube closure,^40^ neurogenesis,^41^ and neuroinflammation.^42^ DNAm of the *BRD2* promotor has been implicated in juvenile myoclonic epilepsy, a common adolescent-onset genetic generalized epilepsy syndrome.^43^ However, we advise caution when interpreting this specific site because, despite having low variation attributable to heterogeneity across the cohorts, its genome-wide significant association with GPF seems to be driven by one single cohort.

With regards to genes with probes at suggestive significance at school-age (*WDR20, MOV10*, and *TAOK2)*, these have previously been linked to neurodevelopmental and psychiatric risk, such as autism spectrum disorder (ASD) and schizophrenia.^44-51^ Pleiotropy was supported by our cross-sectional GWAS enrichment analyses for EWAS, showing that *TAOK2* overlapped with genomic loci previously linked to schizophrenia,^16, 37^ as well as obsessive compulsive disorder and bipolar disorder.^16^ However, despite these previously established links with mental health outcomes, annotated genes of our overall top hits identified in the EWAS meta-analyses were not enriched for common biological processes, cellular components, molecular functions, or pathways.

The significant DMR identified at school-age included 6 CpGs mapped to the *SHC4* gene that is expressed in neurons and regulates *BDNF*-induced MAPK activation. *SHC4* has been associated with multiple types of psychiatric disorders in genome-wide association studies in adulthood, including obsessive-compulsive disorder^52^ and MDD.^53, 54^ The robustness of this association with GPF should be examined mechanistically to understand potential biological processes that may be implicated in patterns shared across psychiatric outcomes in childhood.

Of interest, despite similar sample sizes and measures (i.e., almost exclusively the CBCL), the top signals were very different between the prospective and cross-sectional EWASs, as evidenced for example by the lack of a correlation between nominally significant sites for these analyses. This low overlap might be due to the temporally dynamic nature of the methylome. DNAm patterns vary substantially over time^55^ and can show temporally specific associations with outcomes, including psychiatric symptoms.^20^ Unlike an existing EWAS meta-analysis on ADHD symptoms, which showed the strongest signal prospectively at birth compared to childhood,^20^ we did not detect any significant prospective associations. This is particularly interesting given the use of largely overlapping samples, suggesting that cord blood DNAm may capture risk for specific psychiatric problems (in this case ADHD) rather than a broader liability to psychopathology.

Strengths of this study include the large sample size and the use of DNAm at two different time points (birth and childhood), enabling the assessment of both prospective and cross-sectional associations with GPF. Another important strength is the use of standardized protocols and scripts to fit GPF to the data in a multi-cohort setting. The GPF scores we analyzed were previously found to associate with a module of co-methylated DNAm probes across the genome,^23^ suggesting that it is possible to detect biological correlates of GPF using this study’s measure. Furthermore, the current study showed that GPF consistently negatively correlated with child cognition across the cohorts as expected based on existing evidence,^7^ suggesting that it is capturing a similar, valid construct across the cohorts.

However, the current findings should also be interpreted in the context of several limitations. First, given the possibility of residual confounding and reverse causality, the direction of the observed associations cannot be inferred. DNAm might be a marker for unmeasured environmental factors that could affect GPF via independent pathways.

Furthermore, children with higher levels of mental health problems may evoke a particular environment,^56^ which might affect DNAm. Second, our top hits were unrelated to eQTMs. Future research integrating transcriptomic data will be important for assessing the functional relevance of DNAm changes to gene expression in the brain. Third, because DNAm is tissue specific, our observations in peripheral blood may not reflect DNAm levels in other, potentially more relevant, tissues such as the brain. Finally, despite potential sex differences in mental health problems,^57^ the current study did not examine sex-specificity for power reasons. Future genome-wide studies with larger sample sizes are needed to replicate our findings and further characterize DNAm sites associated with GPF.

In summary, this large EWAS meta-analysis identified one probe (Cg11945228) for which lower DNAm in childhood was associated with higher levels of GPF. Furthermore, one DMR in childhood was associated with GPF. This DMR included 6 CpGs mapped to the *SHC4* gene that has previously been implicated in multiple types of psychiatric disorders in adulthood. In contrast, no prospective associations were identified with DNAm patterns at birth. The current findings call for a more integrative approach to the study of GPF, using multiple omics sources, including the genome, epigenome, and transcriptome, to achieve a more comprehensive understanding of its biological underpinnings.

## Supporting information

Funding and acknowledgements

## Data Availability

Site-level meta-analytical results will be made publicly available upon acceptance for publication. For access to cohort-level data, requests can be sent directly to individual studies.

## Data availability

Site-level meta-analytical results will be made publicly available (https://doi.org/) upon acceptance for publication. For access to cohort-level data, requests can be sent directly to individual studies.

## Code availability

Analytical codes can be requested from authors.

## Acknowledgements

Acknowledgements for each of the participating studies are listed in the Funding and Acknowledgements supplement.

## Compliance with ethical standards

### Ethics

All studies acquired approval from local ethics committees and informed consent was obtained for all participants. Full details are listed in the methods supplement.

