## Supplementary material for "DNA methylation and general psychopathology in childhood: An epigenome-wide meta-analysis from the PACE consortium": Methods supplement

**SUPPLEMENTARY METHODS**

**Supplementary material:** 1
**SUPPLEMENTARY METHODS** 1
*1. Analytical models* 2
*2. Avon Longitudinal Study of Parents and Children (ALSPAC)* 2
*3. Drakenstein Child Health Study (DCHS)* 5
*4. Generation R Study (Generation R)* *8*
*5. Glycyrrhizin in Licorice (GLAKU)* *9*
*6. Human Early Life Exposome (HELIX)* *12*
*7. INfancia y Medio Ambiente (INMA)* 16

### 1. *Analytical models*

***Prospective model*: *DNA methylation at birth* and the general psychopathology factor**

| Outcome | Exposure | Covariates |
|---|---|---|
| General psychopathology factor | DNA methylation at birth | Covariates: maternal smoking status, gestational age, sex, child age at outcome assessment, estimated cell proportions (*Gervin et al., 2019*), batch (optional), maternal age at birth, maternal educational level, ancestry (optional), selection factors (optional) |

***Cross-sectional model*: *Childhood DNA methylation* and the general psychopathology factor**

| Outcome | Exposure | Covariates |
|---|---|---|
| General psychopathology factor | DNA methylation at childhood | Covariates: maternal smoking status, gestational age, sex, child age at outcome assessment, estimated cell proportions (*Houseman et al*., 2012), batch (optional), maternal age at birth, maternal educational level, ancestry (optional), selection factors (optional) |

### 2. *Avon Longitudinal Study of Parents and Children (ALSPAC)*

**Design and study population:** Pregnant women resident in Avon, UK with expected dates of delivery 1st April 1991 to 31st December 1992 were invited to take part in the study (Boyd et al., 2013; Fraser et al., 2013). The initial number of pregnancies enrolled was 14,541 (for these at least one questionnaire has been returned or a "Children in Focus" clinic had been attended by 19/07/99). Of these initial pregnancies, there was a total of 14,676 foetuses, resulting in 14,062 live births and 13,988 children who were alive at 1 year of age. When the oldest children were approximately 7 years of age, an attempt was made to bolster the initial sample with eligible cases who had failed to join the study originally. As a result, when considering variables collected from the age of seven onwards (and potentially abstracted from obstetric notes) there are data available for more than the 14,541 pregnancies mentioned above. The number of new pregnancies not in the initial sample (known as Phase I enrolment) that are currently represented on the built files and reflecting enrolment status at the age of 24 is 913 (456, 262 and 195 recruited during Phases II, III and IV respectively), resulting in an additional 913 children being enrolled. The phases of enrolment are described in more detail in the cohort profile paper and its update (Boyd et al., 2013; Fraser et al., 2013; Northstone et al., 2019). The total sample size for analyses using any data collected after the age of seven is therefore 15,454 pregnancies, resulting in 15,589 foetuses. Of these 14,901 were alive at 1 year of age. A 10% sample of the ALSPAC cohort, known as the Children in Focus (CiF) group, attended clinics at the University of Bristol at various time intervals between 4 to 61 months

of age. The CiF group were chosen at random from the last 6 months of ALSPAC births (1432 families attended at least one clinic). Excluded were those mothers who had moved out of the area or were lost to follow-up, and those partaking in another study of infant development in Avon.
Please note that the study website contains details of all the data that is available through a fully searchable data dictionary and variable search tool" and reference the following webpage: http://www.bristol.ac.uk/alspac/researchers/our-data/.

**Consent and ethical approval:** Ethical approval for the study was obtained from the ALSPAC Ethics and Law Committee and the Local Research Ethics Committees.

**DNA methylation measurements:** As part of the Accessible Resources for Integrated Epigenomic Studies (ARIES, http://www.ariesepigenomics.org.uk/) project, DNA methylation was generated for 1018 mother-offspring pairs from the ALSPAC cohort, using the Infinium HumanMethylation450 BeadChip array (Illumina Inc., San Diego, United States). ARIES participants were selected based on availability of DNA samples at two time points for the mother (antenatal and at follow-up when the offspring were adolescents) and at three time points for the offspring (neonatal, childhood (age 7), and adolescence (age 17)). The current study used child cord blood at birth and whole blood at age 7.

**Generation of methylation data and pre-processing methods**: Methods for methylation measurements in ALSPAC have been described previously (Relton et al., 2015). Briefly, cord blood was collected according to standard procedures. DNA methylation assays and data pre-processing was performed at the University of Bristol as part of the ARIES project. DNA was extracted using standard protocol and was bisulfite-converted using the Zymo EZ DNA MethylationTM kit (Zymo, Irvine, CA). DNA methylation was then measured using the Infinium HM450 BeadChip assay (Illumina Inc, San Diego, CA), according to the standard protocol. Arrays were scanned using an Illumina iScan. An initial review of data quality was assessed using GenomeStudio (version 2011.1). A semi-random approach (sampling criteria were in place to ensure that all time points were represented on each array) was used to distribute ARIES samples across slides to minimize the possibility of potential confounding by batch. Data were normalised using the meffil R package (Min, Hemani, Davey Smith, Relton, & Suderman, 2018) using the functional normalisation approach.

**Child psychopathology data:** Offspring psychopathology was assessed using the parent version of the Development and Well-being Assessment (DAWBA; R. Goodman, Ford, Richards, Gatward, & Meltzer, 2000) at the age of 10 years. The DAWBA band computer prediction variables indicate the probability of disorder in 6 categories, ranging from very unlikely (<0.1%) to probable (>70%). Variables based on the ICD-10 and DSM-IV were used to assess ADHD, Conduct Disorder, Depression, Generalized Anxiety Disorder, Obsessive Compulsive Disorder Social Phobia and Specific Phobia and Oppositional Defiant Disorder (A. Goodman, Heiervang, Collishaw, & Goodman, 2011).The DAWBA band variable for Separation Anxiety was based on the DSM-IV only

**Cell type correction:** Estimated cell type proportion types were obtained using the houseman method (Houseman et al., 2012) with a cord blood reference panel (Gervin et al., 2019) for the prospective analysis and a whole-blood reference panel (Reinius et al., 2012) for the cross-sectional analysis.

**Batch correction:** Batch effects were accounted for by adjusting for 20 surrogate variables, that were generated using the R package SVA (Leek et al., 2019).

**Ancestry/ethnicity:** European ancestry was detected by multidimensional scaling analysis using child GWAS data (Gaunt et al., 2016).

**Smoking during pregnancy:** Maternal smoking during pregnancy was assessed as a categorical variable representing 0 = no smoking during pregnancy, 2 = Stopped before the second trimester of pregnancy and 3 = Smoking in the third trimester or throughout pregnancy.

**Gestational age**: Gestational age was calculated (in days) based on the date of the mother's last menstrual period (LMP) when the mother was certain of this, but for uncertain LMPs and conflicts with clinical assessment the ultrasound assessment was used. Where maternal report and ultrasound assessment conflicted, an experienced obstetrician reviewed clinical records and made a best estimate.

**Child sex:** Offspring biological sex at birth was taken from obstetric records.

**Child age at behavioral assessment**: Child age at the completion of the DAWBA was calculated based on the children's date of birth and the date of DAWBA completion.

**Maternal age**: Continuous (years).

**Maternal education:** Maternal education was assessed in the third trimester of pregnancy and coded as a categorical variable representing: high education = advanced-level school-leaving certificate (post-16)/degree, medium education = ordinary-level school-leaving certificate (at 16) and low education = vocational/certificate of secondary education (at 16, equivalent to lower grades of ordinary-level)/none.

**Child cognition (external variable):** Child IQ was assessed using the WISC-III UK at the age of 8.5 years (Wechsler, Golombok, & Rust, 1992).

**Deviations from analysis plan:** Sample plate correction in ALSPAC led to model convergence errors because of small batches in ALSPAC. Therefore surrogate variable analysis was used to correct for technical variation instead. Further, estimated cell proportions in ARIES cord blood were normalised using the R-package meffil (Suderman, Hemani, & Min, 2019) instead of the FlowSorted.CordBloodCombined.450k R-package (Salas, Gervin, & Jones, 2020).

3. *Drakenstein Child Health Study (DCHS)*

**Design and study population:** Drakenstein Child Health Study (DCHS). The DCHS, a population-based birth cohort,has been described previously (Zar, Barnett, Myer, Stein, & Nicol, 2015). Mothers were enrolled prenatally in their second trimester and followed through pregnancy at two primary care clinics serving two distinct populations (predominantly black African ancestry or predominantly mixed ancestry). Mother-child pairs were followed from birth and infants enrolled in the DCHS were followed until at least five years of age (Zar et al., 2015). All births occurred at a single, central facility, Paarl Hospital. The present study is based on children from the DCHS with DNA methylation data from cord blood, genotyping data, and information on psychopathology factors and covariates.

**Consent and ethical approval:** Ethical approval for human subjects' research was obtained from the Human Research Ethics Committee of the Faculty of Health Sciences of University of Cape Town (HREC UCT REF 401/2009; HREC UCT REF 525/2012). Written informed consent was signed by the mothers on behalf of herself and her infant for participation in this study.

**DNA methylation measurements:** DNA was isolated from cord blood samples that were collected at time of delivery (Morin et al., 2017). DNA methylation was assessed with the Illumina Infinium HumanMethylation450 BeadChips (n=156) and the MethylationEPIC BeadChips (n=160).

Pre-processing and statistics were done using R 3.5.1 (https://www.r-project.org/). Raw iDat files were imported to RStudio where intensity values were converted into beta values. The 450K and EPIC datasets were then combined using the minfi package (Aryee et al., 2014) resulting in 316 samples and 453,093 probes. Background subtraction, color correction and normalization were performed using the preprocessFunnorm function (Fortin et al., 2014).

Samples were determined to be outliers if detected using two or more of the following methods: detectOutlier function from the lumi package (Du, Kibbe, & Lin, 2008), Hannum et al. (2013) method using the locFDR package (https://cran.r-project.org/package=locfdr) and both the outlyx and pfilter functions from the watermelon package (Pidsley et al., 2013). However, no samples we detected in more than one method and so none were removed for this reason. Samples containing maternal blood contamination (n = 33) were removed (Morin et al., 2017). After the completion of pre-processing technical replicates (n = 7) and samples where reported sex did not match sex chromosome methylation signatures (n = 3) were removed leaving a total of 273 samples remaining for downstream analysis.

This dataset contains 59 probes which detect single nucleotide polymorphisms for quality control purposes and so once observed, were removed. Probes with NAs in ≥ 1% of samples or had a detection p value ≥ 1x10-16 in ≥ 1% of samples were removed (n = 10,868). Probes which bind to the sex chromosomes were removed due to the distribution differences observed (n = 9,896). Probes whose sequence contains a SNP either at the CpG site being measured or at the site of the single base pair extension with a minor allele frequency ≥ 1% (Pidsley et al., 2013; Price et al., 2013) were removed (n = 13,598). Autosomal probes which were in silico predicted to non-specifically bind to sex chromosomes in the genome were also removed (n = 9,698) leaving a total of 409,033 probes remaining for downstream analysis (Pidsley et al., 2013; Price et al., 2013).

**Child psychopathology data:** Psychopathology symptoms were assessed with the Child Behavior Checklist 6-18 (CBCL/6-18), a validated and widely used parental assessment of a child's behavioral and emotional problems (Achenbach & Rescorla, 2001). Mothers completed questions about a range of emotional and behavioral problems of the child in the past six months on a three-point scale (0=not true, 1=somewhat true, 2=very true).

**Cell type correction:** Seven default cell types as mentioned in analysis plan. Cord blood cell type composition was predicted using the most recent cord blood reference data set (Gervin et al., 2019) and the IDOL algorithm and probe selection (Koestler et al. 2016).

**Batch correction:** Batch effects were removed using ComBat from the R package sva (Leek et al., 2019).

**Ancestry/ethnicity:** 5 genetic principal components included in the model to adjust for population stratification

**Smoking during pregnancy:** Smoking during pregnancy was assessed based on maternal urine cotinine levels at time of enrollment (second trimester). Passive smokers (exposed in environment but did not smoke themselves, cotinine concentrations >=10 -499 ng/ml) and no smoke exposure (<10 ng/ml) were classified as non-smokers. Active smokers (cotinine concentrations >= 500ng/ml) were classified as smokers. "[0] Non-smokers", "[1] Smokers".

**Gestational age**: Gestational age was recorded from ultrasound measurements in the second trimester of pregnancy. In cases where no ultrasound measurement was available, the expected date of delivery was calculated using symphysis-fundal height, recorded by trained clinical staff at enrolment, or date of last normal menstrual period.

**Child sex:** "[1] female", "[2] male"

**Child age at behavioral assessment**: Continuous (years).

**Maternal age**: Continuous (years).

**Maternal education:** "[0] primary", "[1] some secondary", "[2] completed secondary", "[3] any tertiary"

**Child cognition (external variable):** Child cognitive development was assessed using the Wechsler Preschool and Primary Scale of Intelligence (WPPSI-IV), where cognition was represented by a full-scale IQ (FSIQ) composite score derived from performance across 6 subtests covering verbal and non-verbal areas of cognition; including verbal comprehension, fluid reasoning, visual-spatial ability, processing speed and working memory.

### 4. Generation R Study (Generation R)

**Design and study population:** The Generation R Study is a population-based prospective cohort study (Kooijman et al., 2016). All pregnant women living in Rotterdam, the Netherlands, with an expected delivery date between April 2002 and January 2006 were invited to participate. These women and their children have been followed at regular intervals since recruitment. For the current study, only children of European ancestry were included.

**Consent and ethical approval:** All parents gave informed consent for their children's participation. The Generation R Study is conducted in accordance with the World Medical Association Declaration of Helsinki and study protocols have been approved by the Medical Ethics Committee of the Erasmus Medical Center, Rotterdam.

**DNA methylation measurements:** Preparation and normalization of the Illumina Infinium® HumanMethylation450 BeadChip array data was performed according to the CPACOR workflow using the software package R. In detail, the idat files were read using the *minfi* package. Probes that had a detection p-value above background (based on sum of methylated and unmethylated intensity values) > or equal to 1E-16 were set to missing per array. Next, the intensity values were stratified by autosomal and non-autosomal probes and quantile normalized for each of the six probe type categories separately: type II red/green, type I methylated red/green and type I unmethylated red/green. Beta values were calculated as proportion of methylated intensity value on the sum of methylated+unmethylated+100 intensities. Arrays with observed technical problems such as failed bisulfite conversion, hybridization or extension, as well as arrays with a mismatch between sex of the proband and sex determined by the chromosome X and Y probe intensities were removed from subsequent analyses. Additionally, only arrays with a call rate > 95% per sample were processed further. The final dataset contained information on 458,563 CpGs for 1,396 samples at birth and 464 samples at age 10.

**Child psychopathology data:** Psychopathology symptoms were assessed with the Child Behavior Checklist 6-18 (CBCL/6-18), a validated and widely used parental assessment of a child's behavioral and emotional problems (Achenbach & Rescorla, 2001). Mothers completed questions about a range of emotional and behavioral problems of the child in the past six months on a three-point scale (0=not true, 1=somewhat true, 2=very true).

**Cell type correction:** Cell counts estimated using the Gervin et al. (2019) (DNAm data at birth) or Houseman et al. (2012) (DNAm data at age 10) blood reference panels were included as covariates.

**Batch correction:** Adjustment for batch effects was done by including sample plate as a covariate.

**Ancestry/ethnicity:** Ancestry came from child GWAS data. All children with DNA methylation data were of European ancestry.

**Smoking during pregnancy:** Smoking during pregnancy was assessed with postal questionnaires in early pregnancy (Gestational Age <18 weeks), mid pregnancy (Gestational Age 18-25 weeks) and late pregnancy (Gestational Age>25 weeks). It was classified in three categories: "Never smoked during pregnancy", "Quit when pregnancy was known" and "Continued during pregnancy".

**Gestational age**: (continuous, weeks). Gestational age at birth was established by fetal ultrasound examination

**Child sex:** (0 = female, 1 = male). Child sex was obtained from midwife and hospital registries at birth.

**Child age at behavioral assessment**: Continuous (years).

**Maternal age**: Continuous (years, at intake).

**Maternal education:** Maternal educational level was based on self-reported levels of education during pregnancy. It was classified following the definition of Statistics Netherlands into three-level ordinal categories: 0= high (higher vocational training or higher academic education), 1= medium (>three years general secondary school); 2= low (lower vocational training or three or less years general secondary school).

**Child cognition (external variable):** Child cognition was assessed using a nonverbal IQ test when children were 5–7 years old. Two subtests of the Snijders-Oomen Niet-verbale intelligentie test, 2.5-7- revisie (SON-R 2.5-7; Tellegen, Winkel, Wijnberg-Williams, & Laros, 2005) were administered, including "Mosaics" (spatial insight) and "Categories" (abstract reasoning abilities).

5. Glycyrrhizin in Licorice (GLAKU)

**Design and study population:**

The adolescents of the Glaku (Glycyrrhizin in Licorice) cohort came from an urban community-based cohort comprising 1049 infants of European descent born between March and November 1998 in Helsinki, Finland (Strandberg, Jarvenpaa, Vanhanen, & McKeigue, 2001). In 2009–2011, initial cohort members who had given permission to be contacted and whose addresses were traceable (*N* = 920, 87.7% of the original cohort in 1998) were invited to a follow-up, of which 692 (75.2%) could be contacted by phone (mothers of the adolescents). Of them, 451 (65.2% of those who could be contacted by phone, 49% of the invited) participated in a follow-up at a mean age of 12.3 years (SD = 0.5, range 11.0–13.2 years).

**Consent and ethical approval:** Informed consent was obtained from all participants. The study protocol was approved by the ethical committees of the City of Helsinki and the Uusimaa Hospital District.

**DNA methylation measurements:**

DNA was extracted from child blood samples collected through venepuncture at the mean age of 12.3 years (range 11.1-13.2 years).

DNA was extracted at the National Institute for Health and Welfare, Helsinki, Finland and the Department of Medical and Clinical Genetics, University of Helsinki, Finland [LJMT1] and methylation analyses were performed at the Max Planck Institute in Munich, Germany. DNA was bisulphite-converted using the EZ-96 DNA Methylation kit (Zymo Research). Genome-wide methylation status of over 850 000 CpG sites was measured using the Infinium Methylation EPIC array (Illumina Inc., San Diego, USA) according to the standard protocol in 240 blood samples. The arrays were scanned using the iScan System (Illumina Inc., San Diego, USA). The quality control pipeline was set up using the R-package minfi. Methylation beta-values were normalized using the funnorm function.One IDs showed density artefacts after normalization and was removed from further analysis. We excluded any probes on chromosome X or Y, probes containing SNPs and cross-hybridizing probes according to Chen (Chen et al., 2013), Price (Price et al., 2013) and McCartney (McCartney et al., 2016). Furthermore, any CpGs with a detection p-value > 0.01 in at least 25% of the samples were excluded. The final dataset contains 812,943 CpGs and 239 IDs. We used ComBat to check and adjust for the batch effects.

Genotyping was performed on Illumina Human OmniExpress Exome 1.2 bead chip (Illumina Inc., San Diego, CA) at the Tartu University, Estonia in September 2014 according to the standard protocols. Genomic coverage was extended by imputation using the 1000 Genomes Phase I integrated variant set (v3/April 2012; NCBI build 37/hg19) as the reference sample and IMPUTE2 software. Before imputing the following QC, filters were applied: SNP clustering probability for each genotype > 95%, Call rate > 95% individuals and markers (99% for markers with MAF < 5%), MAF > 1%, HWE p > 1*10–6. Moreover, heterozygosity, sex check, and relatedness checks were performed and any discrepancies were removed (N = 2).

**Child psychopathology data:** Psychopathology symptoms were assessed with the Child Behavior Checklist 6-18 (CBCL/6-18), a validated and widely used parental assessment of a child's behavioral and emotional problems (Achenbach & Rescorla, 2001). Mothers completed questions about a range of emotional and behavioral problems of the child in the past six months on a three-point scale (0=not true, 1=somewhat true, 2=very true).

**Cell type correction:** The Houseman method (Houseman et al., 2012) was applied with Reinius reference data (Reinius et al., 2012) using the estimateCellCounts function from the Minfi package (Jaffe & Irizarry, 2014) in R (https://www.r-project.org/) to estimate the proportions of six white blood cell subtypes (CD4+ T-lymphocytes, CD8+ T-lymphocytes, NK (natural killer) cells, B-lymphocytes, monocytes and granulocytes).

**Batch correction:** We used ComBat to check and adjust for the batch effects.

**Ancestry/ethnicity:** We performed multi-dimensional scaling (MDS) analysis on the identity by state matrix of quality-controlled genotypes. The first three components depicted the origin admixture and were included as covariates in the regression analyses.

**Smoking during pregnancy:** was self-reported and categorized as yes or no.

**Gestational age**: was based on ultrasound scans and derived from the birth register.

**Child sex:** was derived from the Finnish social security number.

**Child age at behavioral assessment**: Continuous (years), calculated from the dates of CBCL assessment and birth date.

**Maternal age**: Continuous (years), derived from the birth register.

**Maternal education:** was assessed by question "What is the highest education level you have achieved" with 8 categories: 1=four-to-eight-year primary school in the former Finnish school system, 2=nine-year primary+lower secondary school in the current Finnish school system, 3=upper secondary education, 4=post-secondary degree, 5=bachelor's degree, 6=master's degree, 7=doctoral dissertation or 8=some other education. There were no occurrences of the categories 1 nor 8 in the data. Since gaining the primary and lower secondary education takes ~9 years in the current Finnish school system, the maternal education level 2 in Glaku would probably correspond to the ISCED levels 0-2 ('low' maternal education in the study plan). However, in the data there were only 4 mothers with the maternal education level 2. Therefore, we combined the maternal education levels 2 and 3. Furthermore, the cumulative years of schooling for upper secondary education, post-degree, bachelor's degree and master's degree are about 12, 14, 15-16, 17-18 and 22, respectively. For the analysis, the categories 2-3 were assigned to 0, categories 4-5 to 1 and categories 6-7 to 2. The resulting frequencies of these categories were 36, 101 and 78, respectively.

**Child cognition (external variable):**
Children were administered two subtests tapping on verbal abilities (Similarities, Vocabulary) and two subtests tapping on non-verbal abilities (Block Design and Picture Arrangement) from the Finnish translation of the Wechsler Intelligence Scales for Children, 3rd Edition (WISC-III). The age and sex standardized scores were summed and converted to z-scores based on the observed distribution to produce estimated IQ.

#### 6. *Human Early Life Exposome (HELIX)*

**Design and study population:** The present study used data from the Human Early Life Exposome Study (HELIX; https://www.projecthelix.eu/), a collaborative project across six established and ongoing longitudinal population-based birth cohort studies in six European countries (Maitre et al., 2018). The project counts with a harmonization protocol for exposures and phenotypes. For this particular analysis, data from six different HELIX cohorts were used: BIB (United Kingdom), EDEN (France), KANC (Lithuania), INMA (Spain), MOBA (Norway) (Magnus et al., 2016), and RHEA (Greece). From the dataset we only included children of European ancestry (selected by genetic background data). The number of participants with both blood DNA methylation (Infinium®HumanMethylation450 BeadChip) and phenotype data (CBCL test) at the age of 7-9 years was 779. Models were adjusted for 20 GWAS PCs to account for genetic substratitification within Europe.

In parallel, the same analysis was conducted with participants of Pakistani ancestry only (selected by genetic background data). In this case, the number of participants with both DNA methylation and phenotype data is 74. For this particular analysis we did not adjust for PCs since all participants are from the same cohort (BIB, United Kingdom).

**Consent and ethical approval:** Prior to the start of HELIX, all six cohorts on which HELIX is based had been in existence for some years, had undergone the required evaluation by national ethics committees and had obtained all the required permissions for their cohort recruitment and follow-up visits. Each cohort also confirmed that relevant informed consent and approval were in place for secondary use of data from pre-existing data. The work in HELIX was covered by new ethics approvals in each country, and at enrolment in the HELIX subcohort and panel studies participants were asked to sign an informed consent form for the specific HELIX work including clinical examination and biospecimen collection and analysis. An Ethics Task Force was established to support the HELIX project on ethical issues, for advice on the project's ethical compliance, identification and alerting to changes in legislation where applicable.

Specific procedures are in place within HELIX to safeguard the privacy of study subjects and confidentiality of data. First, any reported study results pertain to analyses of aggregate data; no variables or combination of variables that can identify an individual will be associated with any published or unpublished report of this study. Primary databases with personal information (such as geocodes, dates, questionnaires or health outcomes) have been stored on separate computers with personal identifiers removed. Subjects are identified by a unique study number, linking all basic data required for the study. The master key file linking the study numbers with personal identifiers is maintained in each cohort. For the dataset analysis, all information that enables identification of an individual (dates, geocodes, etc) is removed before distribution of datasets to the researchers. All data exchanges will adhere to the most up-to-date EU and national data protection regulations.

**DNA methylation measurements:** The following procedure was conducted in the same lab for all the samples from the different cohorts comprising HELIX, which were previously randomized. DNA was obtained from buffy coat collected in EDTA tubes at age 7-9y. Briefly, DNA was extracted using the Chemagen kit (Perkin Elmer) in batches of 12 samples. Samples were extracted by cohort and following their position in the original boxes. DNA concentration was determined in a NanoDrop 1000 UV-Vis Spectrophotometer (ThermoScientific) and with Quant-iT™ PicoGreen™ dsDNA Assay Kit (Life Technologies). DNA methyaltion was assessed with the Infinium HumanMethylation450 beadchip from Illumina, following manufacturer's protocol. Briefly, 700 ng of DNA were bisulfite-converted using the EZ 96-DNA methylation kit following the manufacturer's standard protocol, and DNA methylation measured using the Infinium protocol. A HapMap sample was included in each plate. In addition 24 HELIX inter-plate duplicates were included. Samples were randomized taking into account cohort, sex and panel. Samples from the panel study (samples of the same subject obtained at two time points) were processed in the same array. Two samples were repeated due to their overall low quality. The final number of analyzed samples was 1,361. DNA methylation data were pre-processed using the minfi package (Aryee et al. 2014). Following guidelines of Lehne work (Lehne et al., 2015), we increased the stringency of the detection p-value threshold to 10E-16 and probes not reaching a 98% call rate were excluded. Two samples were filtered due to overall quality: one had a call rate <98% and the other did not pass QC parameters of the MethylAid package (van Iterson et al., 2014). Then, data was normalized with the functional normalization method, which also includes Noob background subtraction and dye-bias correction (Triche, Weisenberger, Van Den Berg, Laird, & Siegmund, 2013). After that, several quality control checks were performed. First, we checked sex consistency using the shinyMethyl package (Fortin et al., 2014) and two samples were excluded. Genetic consistency of duplicates and samples from the same participant was checked with the 450k genotypes. In addition, genetic consistency was evaluated in those samples that had GWAS data and two of them were excluded. Centered-correlation was around 0 for unrelated samples and around 0.8 for duplicates and panel samples. Principal component analysis showed no differential clusters, however a degree of grouping within the cluster was observed for some biological variables (sex, cohort) and for some technical variables. Because of this, we further used COMBAT algorithm (Johnson, Li, & Rabinovic, 2007) to adjust for potential batch effects, using slide as the major known technical bias. Duplicated samples and HapMap samples were removed as well as control probes, probes designed to detect SNPs and probes to measures methylation levels at non-CpG sites. The final dataset for this analysis consisted of 853 HELIX subjects (779 of European ancestry +74 of Pakistani ancestry) with phenotypic data and covariates and 480071 probes.

**Child psychopathology data:** Psychopathology symptoms were assessed with the Child Behavior Checklist 6-18 (CBCL/6-18), a validated and widely used parental assessment of a child's behavioral and emotional problems (Achenbach & Rescorla, 2001). Mothers completed questions about a range of emotional and behavioral problems of the child in the past six months on a three-point scale (0=not true, 1=somewhat true, 2=very true).

**Cell type correction:** blood reference panel with 6 cell types by Houseman et al. (2012).

**Batch correction:** Potential batch effects were adjusted for by the COMBAT algorithm (Johnson et al., 2007), using slide as the major known technical bias.

**Ancestry/ethnicity:** In the analysis with participants of European ancestry, models were adjusted for the first 20 genetic principal components (PCs) to adjust for population stratification. Since the PCs also capture variation due to cohort, we did not adjust for a cohort variable to avoid multicollinearity.

**Smoking during pregnancy:** three-level ordinal category (never smoked during pregnancy, smoked during first trimester only, smoked after first trimester/sustained smoking)

**Gestational age**: Gestational age (continuous, weeks) was established by combination of the variables indicated below and is equal to: - e3_galmp if available, - OR e3_gaultr if e3_galmp not available, - OR e3_gama if e3_galmp and e3_gaultr not available.

e3_galmp: LMP-based GA (date of delivery - date of LMP)/7 (without rounding), where LMP is last menstrual period; e3_galultr: gestational age estimated using ultrasound measurements if there were performed before 20 weeks of gestation: (date of delivery-date of conception estimated by US)/7; e3_gama: GA registered by the maternity records (e3_gama). This is the obstetrician estimation, which is usually based on ultrasound measurements or LMP and possibly corrected for long durations by the obstetrician.

**Child sex:** (0 = female, 1 = male).

**Child age at behavioral assessment**: Continuous (years).

**Maternal age**: Continuous (years).

**Maternal education:** Maternal education was classified into 0 = low (primary or less), 1 = medium (secondary), 2 = high (university).

**Child cognition (external variable):** The total number of correct responses in Raven's Colored Progressive Matrices (Raven & Raven, 1998) was used as a measure of general cognitive index (continuous).

### *7. INfancia y Medio Ambiente (INMA)*

**Design and study population:** The present study used data from participants recruited between 2003 and 2008 in the de novo cohort sited in Sabadell of the INfancia y Medio Ambiente (INMA; http://www.proyectoinma.org/) Project, a population-based mother–child cohort study in Spain (Guxens et al., 2012). Cord blood methylation was measured using the Infinium® HumanMethylation450 BeadChip. The number of participants with both DNA methylation and phenotype data is 292.

**Consent and ethical approval:** The study was approved by the Ethics Committee of the reference hospital, and all participants gave their written informed consent.

**DNA methylation measurements:** Cord blood was extracted using the Chemagen kit (Perkin Elmer). DNA concentration was determined by NanoDrop spectrophotometer (Thermo Scientific) and with the Quant-iT PicoGreen dsDNA Assay Kit (Life Technologies). Methylation data was produced in two different laboratories as part of two different projects: in the Genome Analysis Facility of the University Medical Center Groningen (UMCG) in Holland, and in the Bellvitge Biomedical Research Institute (IDIBELL, Barcelona). Both laboratories used the recommended Illumina protocol for the Infinium HumanMethylation450 beadchip. Briefly, 500 ng of DNA was bisulfite-converted using the EZ 96-DNA methylation kit following the manufacturer's standard protocol, and DNA methylation measured using the Illumina Infinium HumanMethylation450 beadchip.DNA methylation data were preprocessed using the minfi package (Aryee et al., 2014).

A series of steps were completed for quality control and data analysis. The first step was low quality sample removal. First, 2 samples with bad overall quality or with low detection p-value according to the output of the MethylAid package (van Iterson et al., 2014) were removed. Then, we removed 3 samples whose sex was wrongly predicted using shinyMethyl (Fortin et al., 2014). Following guidelines of Lehne work (Lehne et al., 2015), we increased the stringency of the detection p-value threshold to 10-16 and we filtered 18 samples with a call rate lower than 98%. The second step was normalizing data with functional normalization. Correlation between SNP in replicates samples was checked and probes not measuring SNPs were discarded. 7,136 probes with a call rate lower than 95% were also removed. Probes in sexual chromosomes, crosshibridizing or containing SNPs were flagged but not removed at this point. ComBat was applied to remove batch effect (Johnson et al., 2007). Finally, duplicated samples were removed. The final dataset consisted of 292 samples and 476,946 probes with behavioral data available.

**Child psychopathology data:** Psychopathology symptoms were assessed with the Child Behavior Checklist 6-18 (CBCL/6-18), a validated and widely used parental assessment of a child's behavioral and emotional problems (Achenbach & Rescorla, 2001). Mothers completed questions about a range of emotional and behavioral problems of the child in the past six months on a three-point scale (0=not true, 1=somewhat true, 2=very true).

**Cell type correction:** Cord blood reference panel with 7 cell types by Gervin et al. (2019).

**Batch correction:** ComBat was applied to remove batch effect (Johnson et al., 2007).

**Ancestry/ethnicity:** All individuals are classified as Europeans taking into account ethnic origin and country of origin of both parents

**Smoking during pregnancy:** Three-level ordinal category (never smoked during pregnancy, smoked during first trimester only, smoked after first trimester/sustained smoking).

**Gestational age**: Gestational age at blood sampling was calculated based on last menstrual period (LMP) reported at recruitment and confirmed using estimates based on ultrasound examina-tion in the 12th week of gestation. When the difference between the LMP reported at recruitment and estimated from the ultra-sound was ≥ 7 days (n=91; 16%), we esti-mated LMP using a quadratic regression formula (Westerway, Davison, & Cowell, 2000).

**Child sex:** (0 = female, 1 = male). Derived from medical/maternity records at birth

**Child age at behavioral assessment**: Continuous (years).

**Maternal age**: Continuous (years).

**Maternal education:** Maternal education was classified into 0 = low (primary or less), 1 = medium (secondary), 2 = high (university).

**Child cognition (external variable):** General cognitive index (continuous) was assessed using the McCarthy Scales of Children's Abilities (MCSA; McCarthy, 1996) at age 5 (mean=5.09; SD=0.69; Range=4.03-6.86).
