## Supplementary material for "DNA methylation and general psychopathology in childhood: An epigenome-wide meta-analysis from the PACE consortium": Funding and acknowledgements

**FUNDING AND ACKNOWLEDGEMENTS**

### Contents

**Supplementary material:** 1

**FUNDING AND ACKNOWLEDGEMENTS** 1

1. Meta-analysts 2
2. Avon Longitudinal Study of Parents and Children (ALSPAC) 2
3. Drakenstein Child Health Study (DCHS) 3
4. Generation R Study (Generation R) 4
5. Glycyrrhizin in Licorice (GLAKU) 5
6. Human Early Life Exposome (HELIX) 6
7. INfancia y Medio Ambiente (INMA) 8

#### 1. Meta-analysts

**Funding**: The work of Jolien Rijlaarsdam was supported by the Netherlands Organization for Scientific Research (NWO ZonMw VENI, grant no 91618147). The work of Marta Cosin Tomas was supported by a Juan de la Cierva – Formación Postdoctoral Contract awarded by Ministry of Science and Innovation (FJC2018-036335-I), and by a Beatriu de Pinós Postdoctoral Contract awarded by Generalitat de Catalunya-AGAUR and European Commission- Horizon 2020 (2019 BP 00107).

**Role of funders:** The funders had no role in the planning or execution of the study nor the interpretation or publication of its results.

**Acknowledgements**: We are very grateful to the research staff in all of the participating cohorts who contributed to this study. We want to express our deepest gratitude to the thousands of children and their parents who, by participating in the studies in their respective countries, permitted us to conduct this meta-analysis.

#### 2. Avon Longitudinal Study of Parents and Children (ALSPAC)

**Co-authors and affiliations:**

Laura Schellhas, PhD.
School of Psychological Science, MRC Integrative Epidemiology Unit, University of Bristol, Bristol, UK
Institute for Sex Research, Sexual Medicine and Forensic Psychiatry, University Medical Center Hamburg-Eppendorf, Germany

ORCID: 0000-0001-6436-4821

Doretta Caramaschi, PhD
Medical Research Council Integrative Epidemiology Unit, Population Health Science, Bristol Medical School, University of Bristol, Bristol, United Kingdom
Department of Psychology, University of Exeter, Exeter, UK

ORCID: 0000-0002-9740-871X

**Funding**: The UK Medical Research Council and Wellcome (grant ref: 217065/Z/19/Z) and the University of Bristol provide core support for ALSPAC. This publication is the work of the authors and
LS and DC will serve as guarantors for the contents of this paper. Methylation data in the ALSPAC cohort were generated as part of the UK BBSRC funded (grant numbers: BB/I025751/1 and BB/I025263/1) Accessible Resource for Integrated Epigenomic Studies (ARIES, http://www.ariesepigenomics.org.uk). D.C., work in a Unit that is supported by the University of Bristol and the UK Medical Research Council (grant number: MC_UU_00011/5).

**Role of funders:** The funders had no role in the planning or execution of the study nor the interpretation or publication of its results.

**Acknowledgements**: We are extremely grateful to all the families who took part in this study, the midwives for their help in recruiting them, and the whole ALSPAC team, which includes interviewers, computer and laboratory technicians, clerical workers, research scientists, volunteers, managers, receptionists and nurses. We are thankful to Dr Matt Suderman for providing support with the analyses.

**Declaration of competing interests:** None

#### *3. Drakenstein Child Health Study (DCHS)*

**Co-authors and affiliations:**

Anke Huels, PhD
Department of Epidemiology, Rollins School of Public Health, Emory University, Atlanta, GA, USA
Gangarosa Department of Environmental Health, Rollins School of Public Health, Emory University, Atlanta, GA, USA

ORCID: 0000-0002-6005-417X

Sarina Abrishamcar, BS
Department of Epidemiology, Rollins School of Public Health, Emory University, Atlanta, GA, USA

ORCID: 0000-0001-9478-7374

Heather J Zar, MD, PhD
Department of Paediatrics and Child Health, Red Cross War Memorial Children's Hospital, University of Cape Town, SA
South African Medical Research Council (SAMRC) Unit on Child and Adolescent Health, University of Cape Town, Cape Town, South Africa

ORCID: 0000-0002-9046-759X

Dan J Stein, MD, PhD
Department of Psychiatry and Mental Health, University of Cape Town, Cape Town, South Africa
South African Medical Research Council (SAMRC) Unit on Risk and Resilience in Mental Disorders, Neuroscience Institute, University of Cape Town, Cape Town, South Africa

ORCID: 0000-0001-7218-7810

**Funding**: The Drakenstein Child Health Study was funded by the Bill & Melinda Gates Foundation (OPP 1017641, OPP1017579), Medical Research Council South Africa, and the National Research Foundation South Africa. Additional support for the DNA methylation work was by the Eunice Kennedy Shriver National Institute of Child Health and Human Development of the National Institutes of Health (NICHD) under Award Number R21HD085849, and the Fogarty International Center (FIC). AH was supported by the HERCULES Center (NIEHS P30ES019776). DJS and HJZ are supported by the South African Medical Research Council (SAMRC).

**Role of funders:** The funders had no role in the study design, data collection and analysis, decision to publish, or preparation of manuscript.

**Acknowledgements**: The authors thank the study and clinical staff at Paarl Hospital, Mbekweni and TC Newman clinics, as well as the CEO of Paarl Hospital, and the Western Cape Health Department for their support of the study. The authors thank the families and children who participated in this study. The authors also thank Dr. Michael S. Kobor and his team at the University of British Columbia for the generation, pre-processing and quality control of the DNA methylation data (data generation: Julia L MacIsaac, David TS Lin, Katia E Ramadori; pre-processing/quality control: Nicole Gladish).

**Declaration of competing interests:** None.

#### *4. Generation R Study (Generation R)*

**Co-authors and affiliations:**

Jolien Rijlaarsdam, PhD
Department of Child and Adolescent Psychiatry/ Psychology, Erasmus MC University Medical Center Rotterdam, Rotterdam, the Netherlands

ORCID: 0000-0002-2360-518X

Charlotte A. M. Cecil, PhD
Department of Child and Adolescent Psychiatry/ Psychology, Erasmus MC University Medical Center Rotterdam, Rotterdam, the Netherlands
Department of Epidemiology, Erasmus MC University Medical Center Rotterdam, Rotterdam, the Netherlands
Molecular Epidemiology, Department of Biomedical Data Sciences, Leiden University Medical Center, Leiden, The Netherlands

ORCID: 0000-0002-2389-5922

Janine F. Felix, MD, PhD
The Generation R Study Group, Erasmus MC University Medical Center Rotterdam, Rotterdam, the Netherlands
Department of Pediatrics, Erasmus MC University Medical Center Rotterdam, Rotterdam, the Netherlands

ORCID: 0000-0002-9801-5774

Alexander Neumann, PhD
VIB Center for Molecular Neurology, Antwerp, Belgium

ORCID: 0000-0001-6653-3203

**Funding**: The general design of the Generation R Study is made possible by financial support from the Erasmus Medical Center, Rotterdam, the Erasmus University Rotterdam, the Netherlands Organization for Health Research and Development and the Ministry of Health, Welfare and Sport. The EWAS data was funded by a grant from the Netherlands Genomics Initiative (NGI)/Netherlands Organisation for Scientific Research (NWO) Netherlands Consortium for Healthy Aging (NCHA; project nr. 050-060-810), by funds from the Genetic Laboratory of the Department of Internal Medicine, Erasmus MC, and by a grant from the National Institute of Child and Human Development (R01HD068437). The work of Jolien Rijlaarsdam was supported by the Netherlands Organization for Scientific Research (NWO ZonMw VENI, grant no 91618147). The work of Charlotte A M Cecil was funded by the European Union's Horizon 2020 research and innovation programme under the Marie Skłodowska-Curie grant agreement No 707404 and grant agreement No 848158 (EarlyCause). This project received funding from the European Union's Horizon 2020 research and innovation programme (733206, LIFECYCLE; 824989, EUCAN-Connect).

**Role of funders:** The funders had no role in the planning or execution of the study nor the interpretation or publication of its results.

**Acknowledgements:** The Generation R Study is conducted by Erasmus MC in close collaboration with the Faculty of Social Sciences of the Erasmus University Rotterdam, the Municipal Health Service Rotterdam area, the Rotterdam Homecare Foundation, and the Stichting Trombosedienst & Artsenlaboratorium Rijnmond (STAR-MDC), Rotterdam. We gratefully acknowledge the contribution of children and parents, general practitioners, hospitals, midwives and pharmacies in Rotterdam. The generation and management of the Illumina 450K methylation array data (EWAS data) for the Generation R Study was executed by the Human Genotyping Facility of the Genetic Laboratory of the Department of Internal Medicine, Erasmus MC, the Netherlands. We thank Mr. Michael Verbiest, Ms. Mila Jhamai, Ms. Sarah Higgins, Mr. Marijn Verkerk and Dr. Lisette Stolk for their help in creating the EWAS database. We thank Dr. A.Teumer for his work on the quality control and normalization scripts.

**Declaration of competing interests:** None.

#### *5. Glycyrrhizin in Licorice (GLAKU)*

**Co-authors and affiliations:**

Anni Malmberg, MSc,
Department of Psychology & Logopedics, University of Helsinki, Finland,

ORCID: 0000-0003-4347-1788

Kati Heinonen, PhD
Psychology/ Welfare Sciences, Faculty of Social Sciences, Tampere University, Finland
Department of Psychology & Logopedics, University of Helsinki, Finland

ORCID: 0000-0002-1262-5599

Katri Räikkönen, PhD
Department of Psychology & Logopedics, University of Helsinki, Finland

ORCID: 0000-0003-3124-3470

Jari Lahti, PhD
Department of Psychology & Logopedics, University of Helsinki, Finland

ORCID: 0000-0002-4310-5297

**Funding**: The study has been supported by Academy of Finland, University of Helsinki, Hope and Optimism Initiative, Finnish Foundation for Pediatric Research, Sigrid Juselius Foundation, Jalmari and Rauha Ahokas Foundation, Signe and Ane Gyllenberg Foundation, Yrjo Jahnsson Foundation, Juho Vainio Foundation, Emil Aaltonen Foundation, and Ministry of Education and Culture, Finland. The 352 samples were genotyped at the Genotyping and Sequencing Core Facility of the Estonian Genome Centre, University of Tartu.

**Role of funders:** The funders had no role in the planning or execution of the study nor the interpretation or publication of its results.

**Acknowledgements:** We thank all the Glaku children and their parents for their enthusiastic participation. We also thank all the research nurses, research assistants, and laboratory personnel involved in the Glaku study.

**Declaration of competing interests:** None

#### *6. Human Early Life Exposome (HELIX)*

**Co-authors and affiliations:**

Kristine B. Gutzkow, PhD.
Department of Environmental Health, Norwegian Institute of Public Health (NIPH), Oslo, Norway.

ORCID: 0000-0002-6716-5921

Regina Grazuleviciene, MD, PhD
Department of Environmental Science, Vytautas Magnus University, 44248 Kaunas, Lithuania;

ORCID: 0000-0002-0210-8053

John Wright, PhD
Bradford Institute for Health Research, Bradford Teaching Hospitals NHS Foundation Trust, Bradford, UK.

ORCID: 0000-0001-9572-7293

Mariza Kampouri, MSc
Department of Social Medicine, University of Crete, Greece.


Marta Cosin
ISGlobal, Barcelona Institute for Global Health, Barcelona, Spain
Universitat Pompeu Fabra, Barcelona, Spain
Centro de investigación biomédica en red en epidemiología y salud pública (ciberesp), Madrid, Spain

ORCID: 0000-0001-5012-8983

**Funding**: The research leading to these results has received funding from the European Community's Seventh Framework Programme (FP7/2007-206) under grant agreement no 308333—the HELIX project. INMA data collections were supported by grants from the Instituto de Salud Carlos III, CIBERESP, the Conselleria de Sanitat, Generalitat Valenciana, Department of Health of the Basque Government; the Provincial Government of Gipuzkoa, and the Generalitat de Catalunya-CIRIT. KANC was funded by the grant of the Lithuanian Agency for Science Innovation and Technology (6-04-2014_31V-66). The Norwegian Mother, Father and Child Cohort Study is supported by the Norwegian Ministry of Health and Care Services and the Ministry of Education and Research. We are grateful to all the participating families in Norway who take part in this on-going cohort study. The Rhea project was financially supported by European projects, and the Greek Ministry of Health (Program of Prevention of Obesity and Neurodevelopmental Disorders in Preschool Children, in Heraklion district, Crete, Greece: 2011–2014; 'Rhea Plus': Primary Prevention Program of Environmental Risk Factors for Reproductive Health, and Child Health: 2012–2015). The work was also supported by MICINN (MTM2015-68140-R) and Centro Nacional de Genotipado-CEGEN-PRB2-ISCIII.

**Role of funders:** The funders had no role in the planning or execution of the study nor the interpretation or publication of its results.

**Acknowledgements:** The authors would like to thank all the participating children, parents, practitioners and researchers in the six countries who took part in this study. The authors would like to thank Sonia Brishoual, Angelique Serre and Michele Grosdenier (Poitiers Biobank, CRB BB-0033-00068, Poitiers, France) for biological sample management and Professor Frederic Millot (Principal Investigator), Elodie Migault, Manuela Boue and Sandy Bertin (Clinical Investigation Center, Inserm CIC1402, CHU de

Poitiers, Poitiers, France) for planning and investigational actions. The authors would like to thank Veronique Ferrand-Rigalleau, Céline Leger and Noella Gorry (CHU de Poitiers, Poitiers, France) for administrative assistance (EDEN). The authors would like to thank Silvia Fochs, Nuria Pey, Cecilia Persavente and Susana Gross for field work, sample management and overall management in INMA. The authors would like to thank Georgia Chalkiadaki and Danai Feida for biological sample management, to Eirini Michalaki, Mariza Kampouri, Anny Kyriklaki and Minas Iakovidis for field study performance and to Maria Fasoulaki for administrative assistance (RHEA). The authors would also like to thank Jorunn Evandt, Ingvild Essen for thorough field work, Heidi Marie Nordheim for biological sample management and the MoBa administrative unit (MoBa).

**Declaration of competing interests:** None

#### *7. INfancia y Medio Ambiente (INMA)*

**Co-authors and affiliations:**

Jordi Sunyer
ISGlobal, Barcelona Institute for Global Health, Barcelona, Spain
Universitat Pompeu Fabra, Barcelona, Spain
Centro de investigación biomédica en red en epidemiología y salud pública (ciberesp), Madrid, Spain
IMIM Parc Salut Mar, Barcelona, Spain

ORCID: 0000-0002-2602-4110

Silvia Alemany
ISGlobal, Barcelona Institute for Global Health, Barcelona, Spain
Universitat Pompeu Fabra, Barcelona, Spain
Centro de investigación biomédica en red en epidemiología y salud pública (ciberesp), Madrid, Spain

ORCID: 0000-0002-7925-6767

Marta Cosin
ISGlobal, Barcelona Institute for Global Health, Barcelona, Spain
Universitat Pompeu Fabra, Barcelona, Spain
Centro de investigación biomédica en red en epidemiología y salud pública (ciberesp), Madrid, Spain

ORCID: 0000-0001-5012-8983

**Funding**: Main funding of the epigenetic studies, and of birth and nine years of age assessments/visits in INMA were grants from Instituto de Salud Carlos III (Red INMA G03/176, CB06/02/0041, CP18/00018, PI041436, PI081151 incl. FEDER funds, PI12/01890 incl. FEDER funds, CP13/00054 incl. FEDER funds),

CIBERESP, Spanish Ministry of Health (FIS-PI04/1436, FIS-PI08/1151 including FEDER funds, FIS-PI11/00610, FIS-FEDER-PI06/0867, FIS-FEDER-PI03-1615), Spanish Ministry of Economy and Competitiveness (SAF2012-32991 incl. FEDER funds), Agence Nationale de Securite Sanitaire de l'Alimentation de l'Environnement et du Travail (1262C0010), Generalitat de Catalunya-CIRIT 1999SGR 00241, Generalitat de Catalunya-AGAUR (2009 SGR 501, 2014 SGR 822), Fundació La marató de TV3 (090430), EU Commission (261357-MeDALL: Mechanisms of the Development of ALLergy, 308333, 603794, and 634453), and European Research Council (268479-BREATHE: BRain dEvelopment and Air polluTion ultrafine particles in scHool childrEn). We acknowledge support from the Spanish Ministry of Science and Innovation and the State Research Agency through the "Centro de Excelencia Severo Ochoa 2019-2023" Program (CEX2018-000806-S), and support from the Generalitat de Catalunya through the CERCA Program.

**Role of funders:** The funders had no role in the planning or execution of the study nor the interpretation or publication of its results.

**Acknowledgements:** INMA researchers would like to thank all the participants for their generous collaboration. A full roster of the INMA Project Investigators can be found at http://www.proyectoinma.org/presentacion-inma/listado-investigadores/en_listado-investigadores.html.

**Declaration of competing interests:** None.
